# SARS-CoV-2 IgG seroprevalence in the Okinawa Main Island and remote islands in Okinawa, Japan, 2020-2021

**DOI:** 10.1101/2022.03.02.22271759

**Authors:** Kenji Mizumoto, Yusuke Shimakawa, Yoshiaki Aizawa, Christian Butcher, Naomi Chibana, Mary Collins, Kohei Kameya, Tae Gyun Kim, Satoshi Koyama, Ryota Matsuyama, Melissa M. Matthews, Tomoari Mori, Tetsuharu Nagamoto, Masashi Narita, Ryosuke Omori, Noriko Shibata, Satoshi Shibata, Souichi Shiiki, Syunichi Takakura, Naoki Toyozato, Hiroyuki Tsuchiya, Matthias Wolf, Shuhei Yokoyama, Sho Yonaha, Yoshihiro Takayama

**Affiliations:** Okinawa Prefecture Epidemiological Statistics and Analysis Committee, Naha-shi, Okinawa, Japan; Molecular Cryo-Electron Microscopy Unit, Okinawa Institute of Science and Technology Graduate University (OIST), Onna-son, Okinawa, Japan; Research Support Division, Occupational Health and Safety, OIST, Onna-son, Okinawa, Japan; Division of Infectious Diseases, Department of Internal Medicine, Okinawa Prefectural Chubu Hospital, Uruma-shi, Okinawa, Japan; Emergency and critical care center, Okinawa Prefectural Chubu Hospital, Uruma-shi, Okinawa, Japan; Division of Infectious Diseases, Department of Internal Medicine, Okinawa Prefectural Nambu Medical Center & Children’s Medical Center, Haebaru, Shimajiri-gun, Okinawa, Japan; Department of Emergency and Critical Care Medicine, Okinawa Prefectural Nambu Medical Center & Children’s Medical Center, Haebaru, Shimajiri-gun, Okinawa, Japan; Division of Infectious Diseases, Naha City Hospital, Naha-shi, Okinawa, Japan; Emergency Department, Okinawa Prefectural Miyako Hospital, Miyako, Okinawa, Japan; Division of Infectious Diseases, Okinawa Prefectural Yaeyama Hospital, Ishigaki, Okinawa, Japan; Public Kumejima Hospital, Shimajiri-gun, Okinawa, Japan; Graduate School of Advanced Integrated Studies in Human Survivability, Kyoto; University Yoshida-Nakaadachi-Cho, Sakyo-Ku, Kyoto-shi, Kyoto, Japan; Hakubi Center for Advanced Research, Kyoto University, Yoshidahonmachi, Sakyo-Ku, Kyoto-shi, Kyoto, Japan; Unité d’Epidémiologie des Maladies Emergentes, Institut Pasteur, 25-28 rue du Docteur Roux, 75724 Paris, France; International Research Center for Medical Sciences, Kumamoto University, Honjo, Chuo-ku, Kumamoto-shi, Kumamoto, Japan; Vaccine Commercialization Center, Gyeongbuk Institute for Bio industry, Andong, Gyeongbuk, Republic of Korea; Graduate School of Biomedical and Health Sciences, Hiroshima University, Hiroshima, Japan; International Institute for Zoonosis Control, Hokkaido University, Sapporo, Hokkaido, Japan; Division of Bacteriology, Department of Microbiology and Immunology, Faculty of Medicine, Tottori University, Yonago, Tottori, Japan; Department of International Health and Medical Anthropology, Institute of Tropical Medicine, Nagasaki University, Nagasaki, Nagasaki, Japan

## Abstract

We estimated the seroprevalence of anti-SARS-COV-2 IgG in different island groups in Okinawa and described its changes over time. A cross-sectional sero-survey was repeated in three distinct periods between July 2020 and February 2021. A total of 2683 serum samples were collected from six referral medical centers, each covering a separate region in Okinawa. Patients who visited the emergency department for any reason and underwent blood collection were eligible for the study. Samples were analyzed using an FDA-authorized two-step enzyme-linked immunosorbent assay (ELISA) protocol. The case detection ratio was computed by dividing the seroprevalence by the attack rate obtained from publicly available surveillance data. In the main island, the seroprevalence was 0.0% (0/392, 95% CI: 0.0-0.9), 0.6% (8/1448, 0.2-1.1), and 1.4% (8/582, 0.6-2.7) at the 1^st^, 2^nd^, and 3^rd^ sero-survey, respectively. In the remote islands, the seroprevalence was 0.0% (0/144, 95% CI: 0.0-2.5) and 1.6% (2/123, 0.2-5.8) at the 2^nd^ and 3^rd^ survey, respectively. The overall case detection ratios at the 3^rd^ survey were 2.7 (95% CI: 1.3-5.3) in the main island and 2.8 (0.7-11.1) in the remote islands. The highest age-specific case detection ratio was observed in people aged 20-29 years (8.3, 95% CI: 3.3-21.4) in the main island and in those aged 50-59 years (14.1, 2.1-92.7) in the remote islands. The low seroprevalence at the latest survey suggested that a large-scale epidemic had not yet occurred in Okinawa by February 2021. The case detection ratios imply that the cumulative number of incident cases in Okinawa should be 2-3 times higher than that reported by routine surveillance. The ratio was particularly high in young people probably due to a frequent asymptomatic/mild COVID-19 disease in this age group. To accurately measure the scale of the COVID-19 epidemic, it is crucially important to conduct a sero-survey targeting the young.

## Introduction

Since the first case of COVID-19 in Japan was reported on February 14, 2020, Okinawa Prefecture has faced a total of six waves of COVID-19 as of January 10, 2022, with a total of 58209 cases and 398 deaths, compared to 395 patients and seven deaths as of July 31, 2020, when the sero-epidemiological survey was initially planned [1]. Okinawa Prefecture consists of a large number of remote islands centered around the Okinawa Main Island in a vast ocean area. Due to limited medical resources in the remote islands, the dynamics of the COVID-19 epidemic, once it spreads, may differ greatly between the Main Island and the remote islands, including the need for transfer of patients to the larger well-resourced islands by air or sea depending on the case load and clinical severity of the patients at hand.

Since most infected individuals remain mild or asymptomatic, it is widely accepted that the volume of unreported cases of COVID-19 is substantial. Previous studies have reported that the proportion of asymptomatic infections among the elderly groups can reach about 20 percent [2-3]. Despite many mild cases, some may progress to severe diseases resulting in an estimated infection fatality ratio of as high as 6.4% in people over 70 years old [4-6]. Given this high unreported proportion and the poor prognosis for a subset of patients, it is essential to repeat serological surveys over time to obtain a complete picture of the disease dynamics and burden caused by COVID-19. The proportion of previously infected people is a useful indicator to assess the unreported and ascertainment bias regarding the number of cases notified through a surveillance system, the true incidence rate of severe disease and mortality, and the degree of herd immunity.

Large-scale sero-epidemiological surveys have been conducted worldwide, including in Spain and Switzerland, since the first wave of the COVID-19 pandemic [7-8]. In Japan, when this sero-epidemiological survey was originally planned for 2020, there were already a few serosurveys that have been conducted. But these were limited to urban settings (Tokyo, Miyagi, and Osaka) [9] or to a single hospital or private company [10-11].

To the best of our knowledge, this is the first sero-epidemiological survey targeting a wide rural population in Japan. The objective was to determine the seroprevalence of SARS-COV-2 infection in different island groups in Okinawa and to capture the increase in the number of infected residents over time using statistical methods.

## Materials and methods

### Study setting

Okinawa Prefecture is located on the southwestern tip of the Japanese archipelago, and it consists of over 160 large and small islands, including about 50 inhabited islands over a vast ocean area. These islands are divided into Okinawa Main Island and geographically distinct smaller archipelagos such as the Miyako Islands, Yaeyama Islands, and others (Fig 1). Of the approximately 1.45 million people in Okinawa Prefecture, the Main Island accounts for about 1.32 million (91%) and the outlying islands for 130,000 (9%).

**Fig 1.**
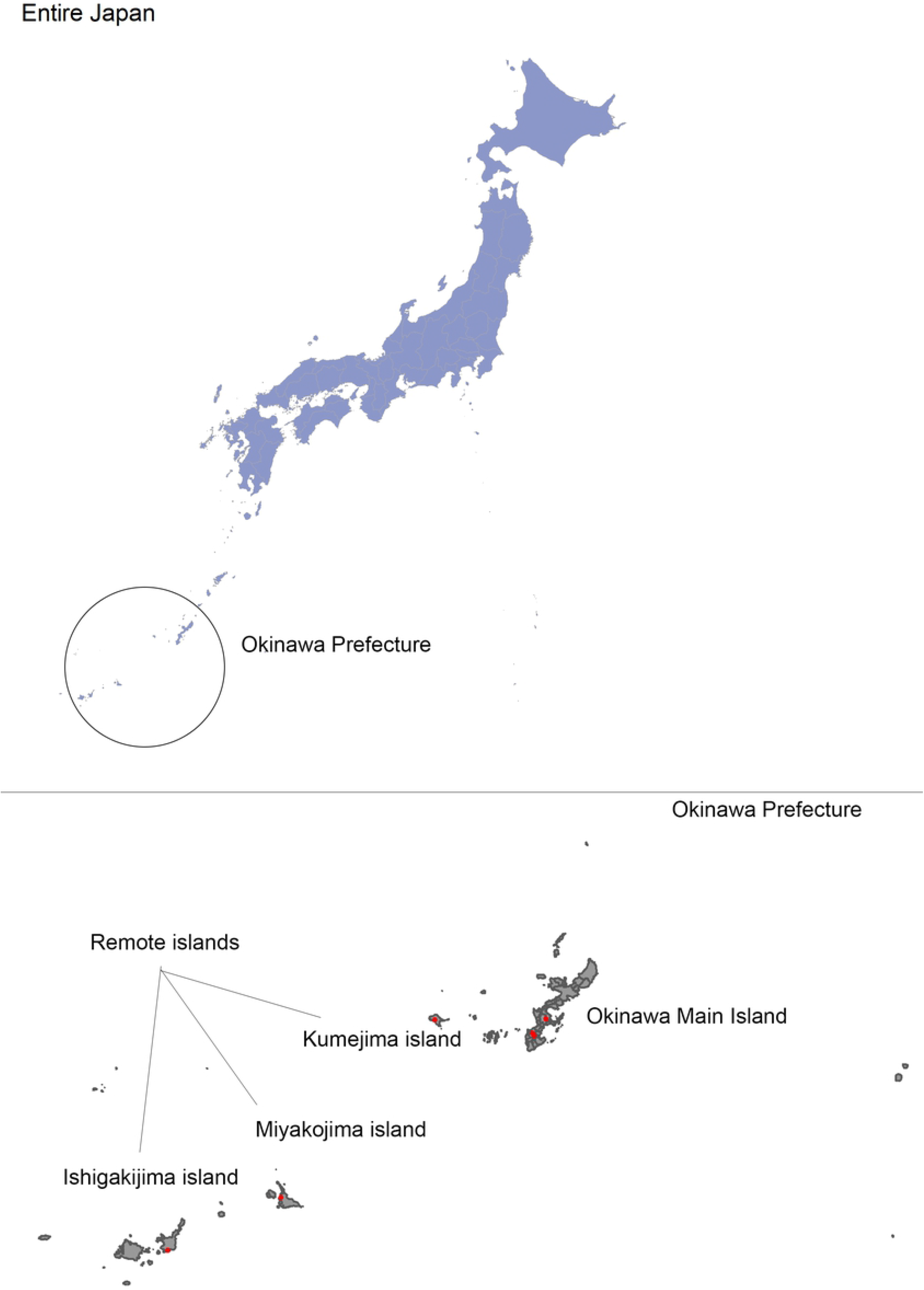
Location of Okinawa Prefecture and six healthcare facilities participated in the study.

Six healthcare facilities participated in the study: Okinawa Prefectural Chubu Hospital (C), Okinawa Prefectural Nambu Medical Center & Children’s Medical Center (NMC), Naha City Hospital (NC), Okinawa Prefectural Miyako Hospital (M), Okinawa Prefectural Yaeyama Hospital (Y), and Public Kumejima Hospital (K). These were selected because they represent a referral medical center located in each region of Okinawa, as presented with red dots in Fig 1. The former three hospitals are tertiary medical institutions located on Okinawa Main Island, and the latter three are secondary hospitals located in Miyako Island, Ishigaki Island, and Kumejima Island, respectively.

### Study design

We conducted a repeated cross-sectional survey in three distinct periods: the 1^st^ survey period was from July 1, 2020, to July 31, 2020, the 2^nd^ survey period was from October 1, 2020, to December 31, 2020, and the 3^rd^ survey period was from February 1, 2021, to February 28, 2021. In remote islands, survey was conducted only in the 2^nd^ and 3^rd^ survey periods. This study was a prospective observational study and carried out on an opt-out basis, with relevant disclosures about the study on the prefectural website and informational posters displayed at each institution. All patients who visited and underwent blood collection for any reason at the emergency departments during these periods were included in this study unless they explicitly chose to be excluded. The remnant leftover serum samples and anonymized demographic data, age group and sex, obtained from medical records were systematically collected. Personal identifiers and identification numbers linking serum samples with medical records were only accessible by a dedicated physician in charge of patient care at each hospital. Serum samples were shipped to Okinawa Institute of Science and Technology Graduate University (OIST), where the serology testing was performed. All the samples were analyzed centrally at OIST using an FDA-authorized 2-step ELISA protocol [12], which detected antibodies against recombinant spike protein purified from mammalian cell culture at OIST.

### Sample size calculation

We defined the sample size for each study period to ensure the precision of an estimate. Assuming a prevalence of 0.5%, recruiting 2,124 people in each period would result in a 95% confidence interval (95%CI) of ±0.3%.

### Antibody measurements

The samples were tested with the FDA-authorized Mount Sinai SARS-CoV-2 2-step ELISA protocol [12], which has a documented sensitivity of 92.5% and specificity of 100% [13]. Briefly, sera diluted 1:50 were tested against SARS-CoV-2 receptor binding domain (RBD). Samples reactive against RBD were tested again by 3-fold serial dilution from 1:100 against full-length, trimeric SARS-CoV-2 spike protein (original Wuhan variant). True positives were defined as samples having signal intensity higher than their plate threshold for both RBD-coated ELISA and for the first two dilutions of the spike-coated ELISA. Plate threshold was defined as 4 times the average of the no serum internal controls (on average 0.19AU).

### Epidemiological data

Serological data collected in C, NMC, and NC were classified as “Okinawa Main Island”, and those collected in M, Y, and K were classified as “remotes islands”. We generated daily time series data of COVID-19 patients for the Okinawa Main Island and remote islands using data obtained from the Okinawa Prefecture about confirmed cases with the date of a confirmed diagnosis, age group, and the municipality of residence, from January 2020 to March 2021. A total of 41 cities, towns, and villages in Okinawa Prefecture are divided into six areas according to the jurisdiction of the public health center: Northern, Central, Southern, Naha City, Miyako, and Yaeyama. In this study, 15 municipalities that are not located on the main island were classified as “remote islands” from a geographical perspective. These are all municipalities under the jurisdiction of the Yaeyama and Miyako Public Health Centers, Ie Village, Iheya Village, and Izena Village under the jurisdiction of the Northern Public Health Center, and Tokashiki Village, Zamami Village, Aguni Village, Tonaki Village, Minami Daito Village, Kita Daito Village, Kumejima Town, and Yaese Town under the jurisdiction of the Southern Public Health Center. The population size by age group and municipality as of January 1, 2020, was obtained from the public source and categorized into Okinawa Main Island and remote islands and by age group [14].

### Statistical analysis

The attack rate was calculated by the number of cumulative confirmed cases divided by the total number of the population. Case detection ratio was defined as a ratio of seroprevalence to attack rate [9]. In order to take into account a latency period between the exposure to the virus and the development of IgG antibodies, we used the attack rate of two weeks before the end of each serosurvey for the calculation of the case detection ratio [15]. Confidence intervals for seroprevalence and attack rates were obtained using the exact binomial test.

All statistical analyses were conducted using R version 4.1.0 (R Foundation for Statistical Computing, Vienna, Austria).

## Results

The number of serum samples collected in the Main Island was 392, 1442, and 582, at the 1^st^, 2^nd,^ and 3^rd^ survey period, respectively. In remote islands, a total of 144 and 123 serum samples were collected at the 2^nd^ and 3^rd^ survey period, respectively. There were no duplicate samples collected from the same individual across the study periods.

Table 1 presented the prevalence of IgG anti-SARS-CoV-2 in Main Island and remote islands for each survey period by age group (0-9, 10-19, 20-29, 30-39, 40-49, 50-59, 60-69, 70-79, and ≥80 years old), by wide age category (0-19, 20-59, ≥60 years old), and by gender. The age-specific seroprevalences at each period are also presented in Fig 2.

**Table 1.**
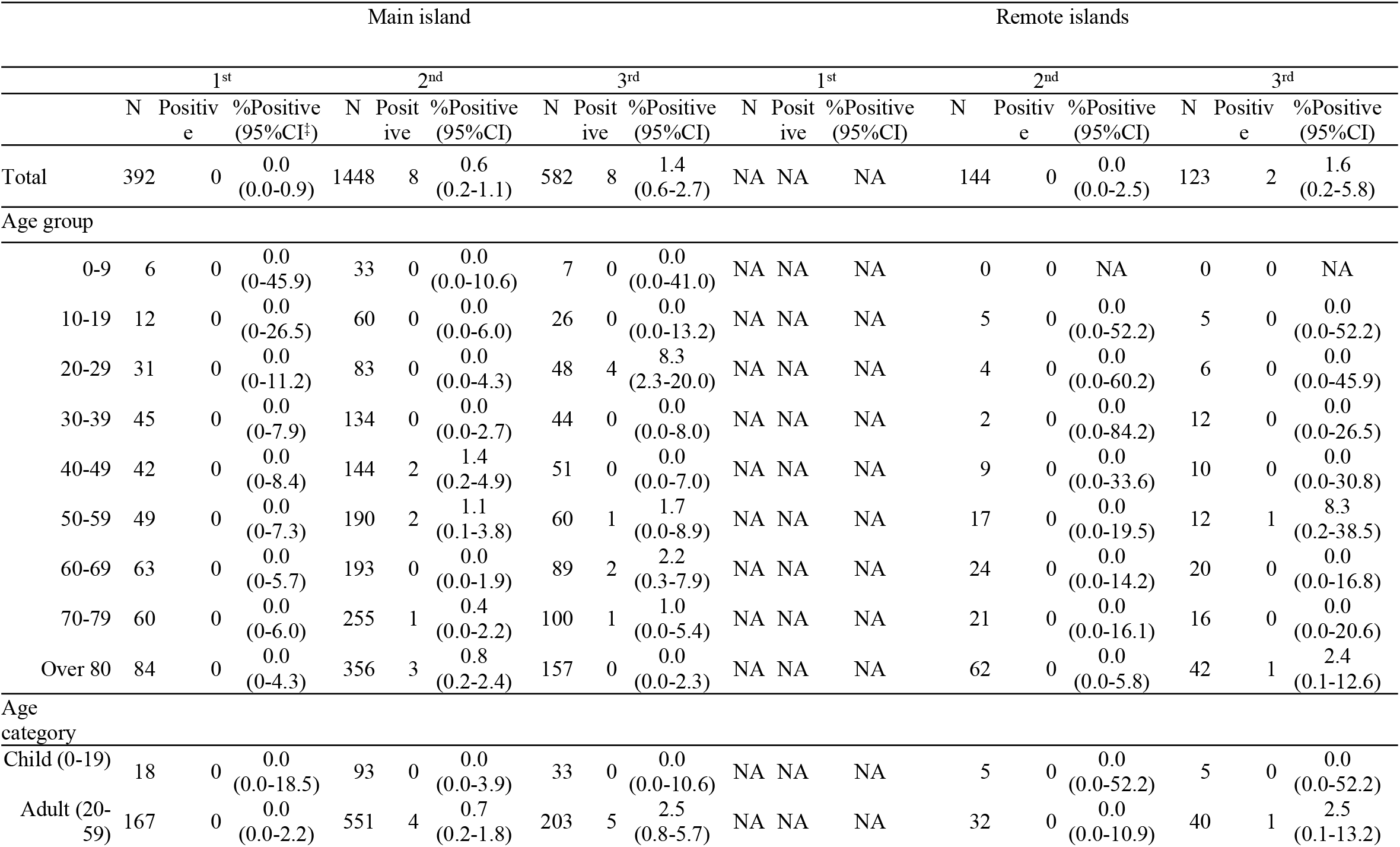
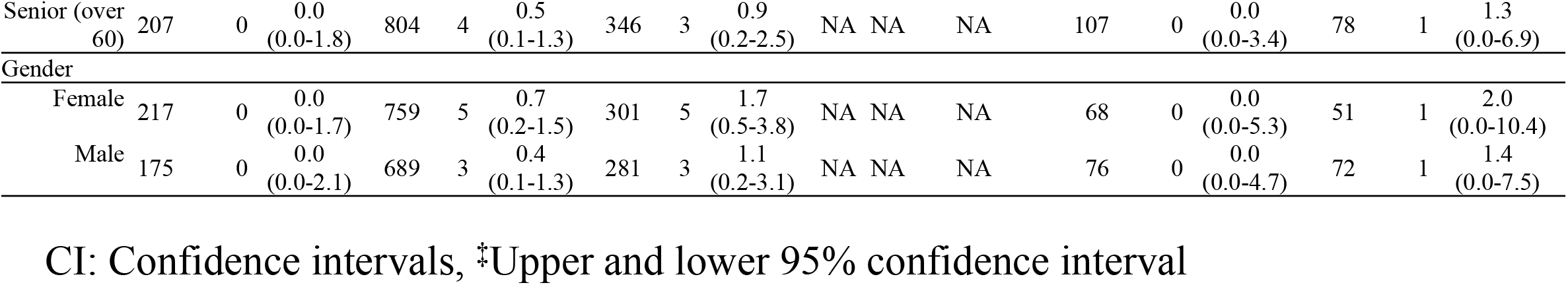
Patient characteristics and the number of positive cases by survey period stratified by regions, Okinawa, 2020-2021.

|  | Main island |  |  |  |  |  |  |  |  | Remote islands |  |  |  |  |  |  |  |  |
| --- | --- | --- | --- | --- | --- | --- | --- | --- | --- | --- | --- | --- | --- | --- | --- | --- | --- | --- |
|  | 1<sup>st</sup> |  |  | 2<sup>nd</sup> |  |  | 3<sup>rd</sup> |  |  | 1<sup>st</sup> |  |  | 2<sup>nd</sup> |  |  | 3<sup>rd</sup> |  |  |
|  | N | Positive | %Positive (95%CI) | N | Positive | %Positive (95%CI) | N | Positive | %Positive (95%CI) | N | Positive | %Positive (95%CI) | N | Positive | %Positive (95%CI) | N | Positive | %Positive (95%CI) |
| Total | 392 | 0 | 0.0 (0.0-0.9) | 1448 | 8 | 0.6 (0.2-1.1) | 582 | 8 | 1.4 (0.6-2.7) | NA | NA | NA | 144 | 0 | 0.0 (0.0-2.5) | 123 | 2 | 1.6 (0.2-5.8) |
| Age group |  |  |  |  |  |  |  |  |  |  |  |  |  |  |  |  |  |  |
| 0-9 | 6 | 0 | 0.0 (0-45.9) | 33 | 0 | 0.0 (0.0-10.6) | 7 | 0 | 0.0 (0.0-41.0) | NA | NA | NA | 0 | 0 | NA | 0 | 0 | NA |
| 10-19 | 12 | 0 | 0.0 (0-26.5) | 60 | 0 | 0.0 (0.0-6.0) | 26 | 0 | 0.0 (0.0-13.2) | NA | NA | NA | 5 | 0 | 0.0 (0.0-52.2) | 5 | 0 | 0.0 (0.0-52.2) |
| 20-29 | 31 | 0 | 0.0 (0-11.2) | 83 | 0 | 0.0 (0.0-4.3) | 48 | 4 | 8.3 (2.3-20.0) | NA | NA | NA | 4 | 0 | 0.0 (0.0-60.2) | 6 | 0 | 0.0 (0.0-45.9) |
| 30-39 | 45 | 0 | 0.0 (0-7.9) | 134 | 0 | 0.0 (0.0-2.7) | 44 | 0 | 0.0 (0.0-8.0) | NA | NA | NA | 2 | 0 | 0.0 (0.0-84.2) | 12 | 0 | 0.0 (0.0-26.5) |
| 40-49 | 42 | 0 | 0.0 (0-8.4) | 144 | 2 | 1.4 (0.2-4.9) | 51 | 0 | 0.0 (0.0-7.0) | NA | NA | NA | 9 | 0 | 0.0 (0.0-33.6) | 10 | 0 | 0.0 (0.0-30.8) |
| 50-59 | 49 | 0 | 0.0 (0-7.3) | 190 | 2 | 1.1 (0.1-3.8) | 60 | 1 | 1.7 (0.0-8.9) | NA | NA | NA | 17 | 0 | 0.0 (0.0-19.5) | 12 | 1 | 8.3 (0.2-38.5) |
| 60-69 | 63 | 0 | 0.0 (0-5.7) | 193 | 0 | 0.0 (0.0-1.9) | 89 | 2 | 2.2 (0.3-7.9) | NA | NA | NA | 24 | 0 | 0.0 (0.0-14.2) | 20 | 0 | 0.0 (0.0-16.8) |
| 70-79 | 60 | 0 | 0.0 (0-6.0) | 255 | 1 | 0.4 (0.0-2.2) | 100 | 1 | 1.0 (0.0-5.4) | NA | NA | NA | 21 | 0 | 0.0 (0.0-16.1) | 16 | 0 | 0.0 (0.0-20.6) |
| Over 80 | 84 | 0 | 0.0 (0-4.3) | 356 | 3 | 0.8 (0.2-2.4) | 157 | 0 | 0.0 (0.0-2.3) | NA | NA | NA | 62 | 0 | 0.0 (0.0-5.8) | 42 | 1 | 2.4 (0.1-12.6) |
| Age category |  |  |  |  |  |  |  |  |  |  |  |  |  |  |  |  |  |  |
| Child (0-19) | 18 | 0 | 0.0 (0.0-18.5) | 93 | 0 | 0.0 (0.0-3.9) | 33 | 0 | 0.0 (0.0-10.6) | NA | NA | NA | 5 | 0 | 0.0 (0.0-52.2) | 5 | 0 | 0.0 (0.0-52.2) |
| Adult (20-59) | 167 | 0 | 0.0 (0.0-2.2) | 551 | 4 | 0.7 (0.2-1.8) | 203 | 5 | 2.5 (0.8-5.7) | NA | NA | NA | 32 | 0 | 0.0 (0.0-10.9) | 40 | 1 | 2.5 (0.1-13.2) |
| Senior (over 60) | 207 | 0 | 0.0<br>(0.0-1.8) | 804 | 4 | 0.5<br>(0.1-1.3) | 346 | 3 | 0.9<br>(0.2-2.5) | NA | NA | NA | 107 | 0 | 0.0<br>(0.0-3.4) | 78 | 1 | 1.3<br>(0.0-6.9) |
| Gender |  |  |  |  |  |  |  |  |  |  |  |  |  |  |  |  |  |  |
| Female | 217 | 0 | 0.0<br>(0.0-1.7) | 759 | 5 | 0.7<br>(0.2-1.5) | 301 | 5 | 1.7<br>(0.5-3.8) | NA | NA | NA | 68 | 0 | 0.0<br>(0.0-5.3) | 51 | 1 | 2.0<br>(0.0-10.4) |
| Male | 175 | 0 | 0.0<br>(0.0-2.1) | 689 | 3 | 0.4<br>(0.1-1.3) | 281 | 3 | 1.1<br>(0.2-3.1) | NA | NA | NA | 76 | 0 | 0.0<br>(0.0-4.7) | 72 | 1 | 1.4<br>(0.0-7.5) |

CI: Confidence intervals, ‡Upper and lower 95% confidence interval

**Fig 2.**
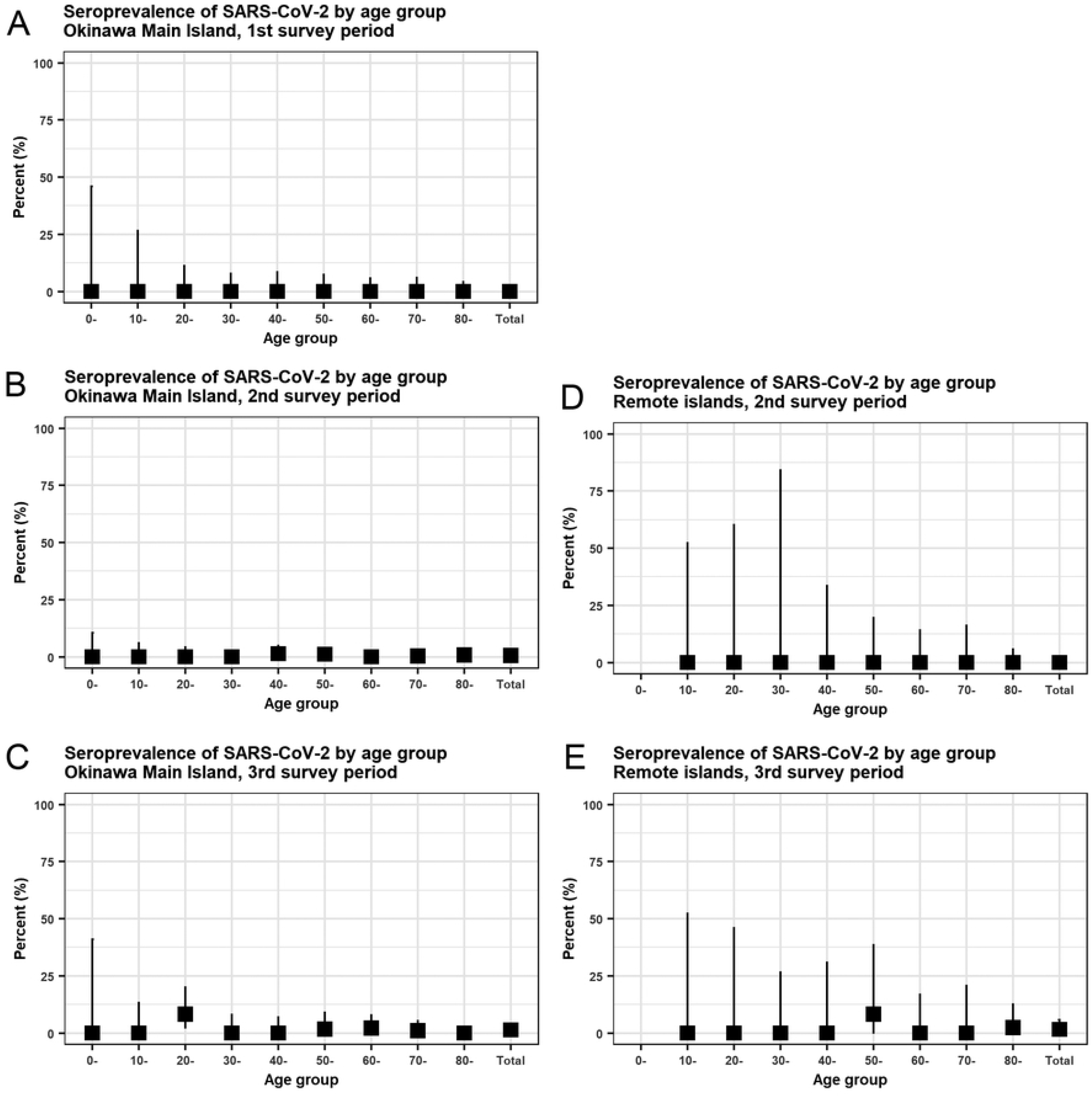
SARS-CoV-2 IgG seroprevalence by age group in Okinawa Main Island and remote islands during the 1^st^, 2^nd^, and 3^rd^ survey periods, Japan, 2020-2021. (A) Main Island, 1^st^ survey period, (B) Main Island, 2^nd^ survey period, (C) Main Island, 3^rd^ survey period, (D) remote islands, 2^nd^ survey period, (E) remote islands, 3^rd^ survey period. There is no available data for under 10 years old in those age in the 2^nd^ and 3^rd^ survey periods in the remote islands.

The seroprevalence in Main Island was 0.0% (0/392, 95%CI: 0.0-0.9%) for the 1^st^ survey period, 0.6% (8/1448, 95%CI: 0.2-1.1%) for the 2^nd^ survey period, and 1.4% (8/582, 95%CI: 0.6-0.7%) for the 3^rd^ survey period. In remote islands the seroprevalence was 0.0% (0/144, 95%CI: 0.0-2.5%) for the 2^nd^ survey period and 1.6% (2/123, 95%CI: 0.2-5.8) for the 3^rd^ survey period.

In Main Island, seroprevalence was taking the average value of zero with different upper 95%CI for all subgroups in age group, age category, and gender for the 1^st^ survey period. For the 2^nd^ survey period, some subgroups were taking the average positive values. They were: those aged 40-49 year-old, 1.4% (95%CI: 0.2-4.9%); those aged 50-59 year-old, 1.1% (95%CI: 0.1-3.8%); those aged 70-79 year-old, 0.4% (95%CI: 0.2-2.2%); and those aged 80 years and above, 0.8% (95%CI: 0.2-2.4%). By age category, it was 0.0% (95%CI: 0.0-3.9%) for children aged 0-19 years, 0.7% (95%CI: 0.2-1.8%) for adults aged 20-59 years, and 0.5% (95%CI: 0.1-1.3%) for seniors aged 60 years and over. By gender, it was 0.7% (95%CI: 0.2-1.5%) for female, and 0.4% (95%CI: 0.1-1.3%) for male.

Finally, for the 3^rd^ survey period, the seroprevalences had seen an increase relative to those in the previous survey periods. The age-specific prevalence was 8.3 (95%CI: 2.3-20.0%) for 20-29 years-old; 1.7 % (95%CI: 0.0-8.9%) for 50-59 years-olds; 2.2 (95%CI: 0.3-7.9%) for 60-69 years-old; and 1.0% (95%CI: 0.0-5.4%) for 70-79 years-old.

By using wider age category, the prevalence was 0.0% (95%CI: 0.0-10.6%) for children, 2.5% (95%CI: 0.8-5.7%) for adults, and 0.9 % (0.2-2.5%) for seniors. The sex-specific prevalence was 1.7% (95%CI: 0.5-3.8%) for female and 1.1% (95%CI: 0.2-3.1%) for male.

In remote islands, seroprevalence was taking the average value of zero with different uncertainty bounds for all subgroups in age group, age category, and gender for the 2^nd^ survey period. For the 3^rd^ survey period, some subgroups were taking the average positive values: 8.3% (95%CI: 0.2-38.5%) for those aged 50-59 year-old and 2.4% (95%CI: 0.1-12.6%) for those aged 80 years and above.

By using wider age category, the prevalence was 0.0% (95%CI: 0.0-52.2%) for children, 2.5% (95%CI: 0.1-13.2%) for adults and 1.3 % (0.0-6.9%) for seniors. The sex-specific prevalence was 2.0% (95%CI: 0.0-10.4%) for female and 1.4% (95%CI: 0.0-7.5%) for male.

Fig 3 presented the attack rates in Main Island and remote islands by age groups at three distinct time points: July 15, 2020; November 15, 2020; and February 15, 2021. These are intermediate time points for corresponding survey periods.

**Fig 3.**
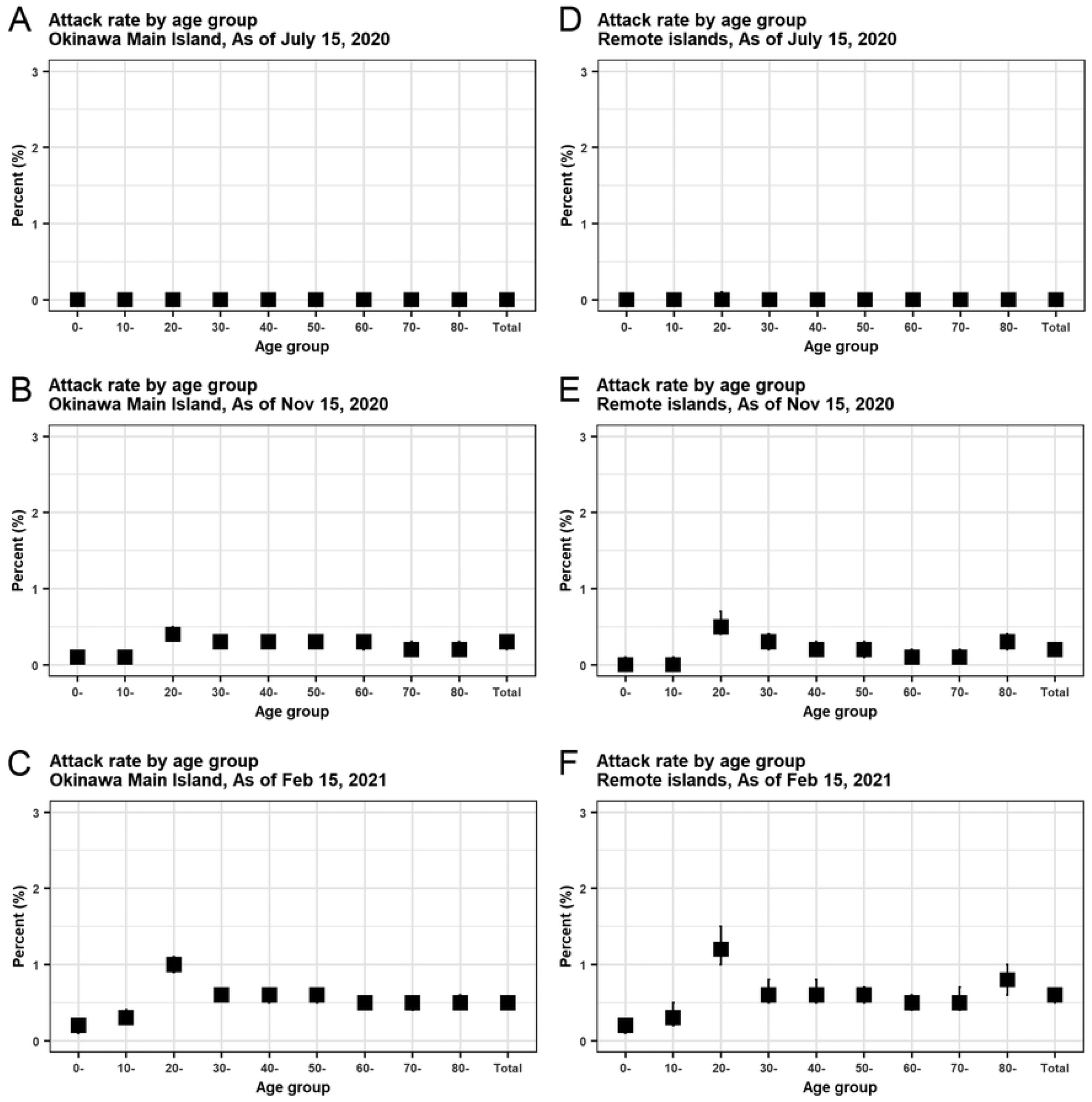
Attack rates of COVID-19 by age group in Okinawa Main Island and remote islands during the 1st, 2nd, and 3rd survey periods, Japan, 2020-2021. (A) Main Island, 1st survey period, (B) Main Island, 2nd survey period, (C) Main Island, 3rd survey period, (D) remote islands, 2nd survey period, (E) remote islands, 3rd survey period.

Temporal distributions of confirmed cases and cumulative confirmed cases in Main Island and remote islands were presented in Fig 4, where three study periods were highlighted in grey.

**Fig 4.**
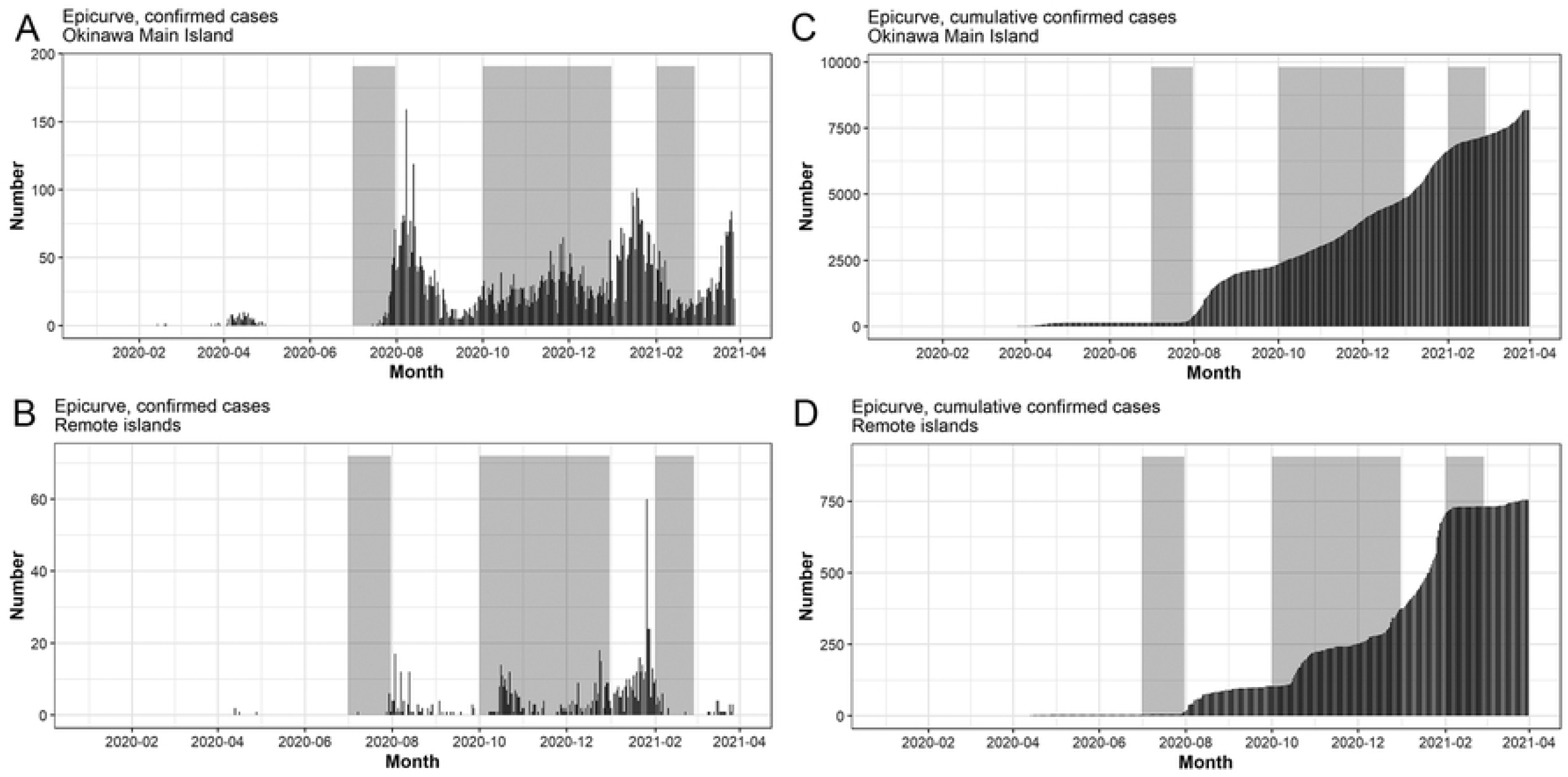
Temporal distribution of COVID-19cases by Okinawa Main Island and remote islands, Okinawa Prefecture in Japan, March 2020-March 2021. Incidence cases in (A) Main Island and (B) Remote islands over time. Cumulative cases in (C) Main Island and (D) Remote islands over time. As the dates of illness onset were not available, we used dates of confirmation.

The overall case detection ratios were 2.2 (95%CI: 1.1-4.4) for the 2^nd^ survey period and 2.7 (95%CI: 1.3-5.3) for the 3^rd^ survey period in Main Island, and 2.8 (95%CI: 0.7-11.1) for the 3^rd^ survey period in remote islands. Due to the insufficient number of seropositive cases, the case detection ratio was not obtained for the 1^st^ survey period in Main Island and 2^nd^ survey period in remote islands. We then investigated which age groups had a relatively high case detection ratio. In the 2^nd^ survey period in the Main Island, the highest and second-highest age-specific case detection ratios were observed in the age group of 40-49 years (4.6, 95%CI: 1.2-18.2), 50-59 years (3.7, 95%CI: 0.9-14.6) and 80-89 years (3.7, 95%CI: 1.2-11.4). In the 3^rd^ survey period in the Main Island, the age-specific case detection ratio was the highest in the age group of 20-29 years (8.3, 95%CI: 3.3-21.4), followed by those aged 60-69 years (4.5, 95%CI: 1.2-17.9). In the 3^rd^ survey period in the remote islands, the ratio was the highest in the age group of 50-59 years (14.1, 95%CI: 2.1-92.7), followed by those aged ≥80 years (3.0, 95%CI: 0.4-21.0).

## Discussion

By repeating sero-epidemiological surveys at three different periods between 2020 and 2021, we obtained the estimates of the proportion of previously infected people in Okinawa Main Island and remote islands. The study findings suggested that the majority of the local population in Okinawa Island were not infected with SARS-CoV-2 as of February 2021. This implies that a large-scale epidemic had not yet occurred. In addition, the estimated seroprevalence among the elderly over 60 years old was relatively low. Although there may have been fewer opportunities for social activities that put the elderly at risk of infection, early communication of the high mortality rate among infected elderly through the media may have led to a behavior of the elderly people and their careers in favor of preventing the infection [4]. In Japan, priority was given to the vaccination of health care workers and then the elderly against the COVID-19. The vaccination of healthcare workers and elderly people started on February 17, 2021, and April 12, 2021, respectively[16]. Given that the last survey period was until February 28, 2021, local population, except for a very small number of health care workers, had not been vaccinated during the study period. Our observation that the most of the survey participants were susceptible to the infection supports the rationale for prioritizing elder population, who are at higher risk of death, for vaccination as a public health policy in Japan.

The seroprevalence of 0.0% (0/392, 95% CI: 0.0-0.9%) in July 2020 in the Okinawa Main Island was quite similar to the seroprevalence of 0.0% (1/3009, 95% CI: 0.0-0.2) in Miyagi Prefecture in early June 2020, which was known to have a relatively low attack rate of 0.004% (88 cases/2.3 million population) as of May 31, 2020 (9). We expected that the seroprevalence in Okinawa in the 1^st^ survey should be higher than that in Miyagi Prefecture for two reasons. First, the attack rate in Okinawa Prefecture as of May 31 was 0.010% (142 cases/1.4 million people), which is about 2.5 times higher than that in Miyagi Prefecture. Second, the study setting in Okinawa’s serosurvey was emergency outpatient, while in Miyagi’s serosurvey this was a random sample from the civil registry. Possible reasons for this unexpectedly low seroprevalence rate in our study in Okinawa include the insufficient sample size in the serosurvey and a potential for selection bias due to a difference in the risk of infection between the general population and those visiting in the emergency departments.

The prefectural-level case detection ratios that ranged from 2.2 to 2.8 in our study were similar to those obtained in other parts of Japan during the same period (2.6-8.7) and considerably lower than those reported in Switzerland (≈20-50) [9, 17]. These results suggest that the surveillance system in Japan may capture the actual number of incident cases better than other countries. The expansion of the PCR testing capacity and monitoring system during a pandemic represents a key intervention, and we believe that the Okinawa Prefecture was moderately well prepared in the PCR screening and monitoring system to grasp the actual status of the epidemic. In the 2^nd^ survey period, when the spread of the epidemic was still limited, the case detection ratios of middle-aged and older people were higher than those in younger age groups. However, in the 3^rd^ survey period, when the epidemic spread more widely, the detection ratios of people in the age group of 20-29 years were much higher than those in other age-groups. Since the vast majority of young people have asymptomatic or mild disease, it is likely that many of them do not come for testing. To get a complete picture of the epidemic, PCR testing and monitoring systems need to be expanded, especially to improve access to testing and care for young people.

In the remote islands, the characteristics of the distribution of the attack rate were similar to those of the Main Island, especially in the 3^rd^ period when the epidemic was widespread. However, because we could not obtain enough samples in the serological survey, the uncertainty bounds were quite wide, and we were not able to obtain precise estimates. More participants would have been needed to accurately estimate the burden of COVID-19. We also had a sample of 33 people in their 50s or younger in the midst of the epidemic, but none were positive. Young people are healthier and less likely to visit emergency departments.

The number of confirmed cases among young people was low in 2020 but increased in early 2021. It has been pointed out that the small number of young people in 2020 may be due to measurement bias, where testing was not conducted in low-risk young people because of insufficient testing systems. However, in the present study, the low serological positivity rate among young people in the early stage of the 2020 outbreak suggests the possibility that the COVID-19 was not actually prevalent among young people, rather than measurement bias.

Our study has limitations. As with other serological studies, our current study is not entirely free of selection bias. The age distribution of the general population and that of healthcare-seeking patients at emergency departments should be quite different. Indeed, the age distribution of participants in the current analysis was skewed towards older people, potentially impacting the overall estimates of seroprevalence.

Furthermore, emergency outpatients may not accurately represent the local population in terms of the risk of exposure to COVID-19 and preventive behaviors, including the uptake of the vaccine. This “unrepresentativeness” may be more prominent in secondary care hospitals, where disease severity is high. The survey period differed somewhat among the six facilities. This may have had some effects on the results.

## Conclusion

In conclusion, this study revealed that the majority of the local population in Okinawa was still susceptible to the COVID-19 disease, indicating that a large-scale epidemic had not yet occurred among residents in Okinawa Prefecture. When the spread of the epidemic materializes, high-intensity countermeasures would be required, focusing primarily on the elderly, who are at high risk of death. Furthermore, it was shown that as the epidemic progresses, the case detection ratio would be higher among younger people, who have a higher proportion of asymptomatic and mild cases and do not actively seek testing. The Omicron variant, which is currently spreading globally, apparently has an even higher rate of mild illness than the previous strains, particularly among young people; this indicates that the number of reported cases might hugely underestimate the current incidence of the infection. In order to accurately measure the scale of the COVID-19 epidemic, it is crucially important to conduct a serological survey targeting the young age group.

## Data Availability

The data that support the findings of this study are available from the corresponding author (Dr. Kenji Mizumoto) and Okinawa Prefecture Government, but restrictions apply to the availability of these data, which were used under license for the current study, and so are not publicly available. Data are however available from the authors upon reasonable request and with permission of Okinawa Prefecture Government.

## List of abbreviations

COVID-19: Coronavirus Disease 2019
RT-PCR: Reverse Transcription Polymerase Chain Reaction
SARS-CoV-2: Severe Acute Respiratory Syndrome Virus 2

## Acknowledgments

Authors appreciate the following institutions for their contribution to support this study: Okinawa Prefecture, Sanitary and Environmental Research Institute in Okinawa Prefecture, Public Health Centers, Public Health Institutes, and medical facilities in Okinawa Prefecture.

## Authors’ contributions

YT conceived and directed the project. KM and YT designed the study with support from YS. KM conducted the analysis and wrote the first draft with support from YS, and TY. NT, RM and RO gave additional feedback on the first draft. YA, MN, SS, ST, SY, NT, HT, NC, SK, KK and SY collected the serological data and gave medical advice. M.M., ELISA implementation, validation, analysis. M.M., S.S., N.S., ELISA execution. T.G.K., N.S., protein expression. T.G.K., protein purification. C.B., workflow automation. M.N. provided samples and controls. M.N. and T.M, medical advice. M.C., advice on study design and ethics. M.W. OIST project creation and supervision. All authors reviewed the results and approved the final version of the manuscript.

## Ethics approval and consent to participate

This study was approved by the ethics committees of Okinawa Prefectural Chubu Hospital (approval number: 24 in 2020), as well as all participating institutions (Okinawa Prefectural Chubu Hospital, Okinawa Prefectural Nambu Medical Center & Children’s Medical Center, Naha City Hospital, Okinawa Prefectural Miyako Hospital, Public Kumejima Hospital, OIST). This study was approved by the ethics committee of each of all six participating institutions. OIST HSR approval number HSR-2020-026.

All methods, including obtaining informed consent, were conducted in accordance with the Declaration of Helsinki and other relevant guidelines, including the Ethical Guidelines for Medical and Health Research Involving Human Subjects set forth by the Japanese government. Consent to publish the accumulated anonymized data has been obtained from all participants of the study upon enrollment.

## Consent for publication

Not applicable.

